# Depletion-of-susceptibles bias in influenza vaccine waning studies: how to ensure robust results

**DOI:** 10.1101/19003616

**Authors:** M. Lipsitch, E. Goldstein, G.T. Ray, B. Fireman

## Abstract

Vaccine effectiveness (VE) studies are subject to biases due to depletion of at-risk persons or of highly susceptible persons at different rates from different groups (depletion-of-susceptibles bias), a problem that can also lead to biased estimates of waning effectiveness, including spurious inference of waning when none exists. An alternative study design to identify waning is to study only vaccinated persons, and compare for each day the incidence in persons with earlier or later dates of vaccination. Prior studies suggested under what conditions this alternative would yield correct estimates of waning. Here we define the depletion-of-susceptibles process formally and show mathematically that for influenza vaccine waning studies, a randomized trial or corresponding observational study that compares incidence at a specific calendar time among individuals vaccinated at different times before the influenza season begins will not be vulnerable depletion-of-susceptibles bias in its inference of waning under the null hypothesis that none exists, and will – if waning does actually occur – underestimate the extent of waning. Such a design is thus robust in the sense that a finding of waning in that inference framework reflects actual waning of vaccine-induced immunity. We recommend such a design for future studies of waning, whether observational or randomized.

---

Recent studies of influenza vaccine effectiveness (VE)have suggested that effectiveness declines over the course of one season, i.e., that vaccine efficacy declines as the season progresses[1-3]. However, these results have been called into question because inferences of waning may be biased. When there is no waning, some study designs (including the classic test-negative observational design [4, 5] and randomized controlled trials [6-8]) may nonetheless infer waning – measured as a decline in vaccine effectiveness as the season progresses. This biased inference is predicted to occur when the vaccine offers “leaky” protection, reducing the probability of infection on exposure by some proportion less than 100%, and either or both of the following conditions holds and is unaccounted for in the analysis [5]: i) some infections occur unobserved in the study population, such that individuals are infected and (for the season) immune to further infection unbeknownst to the researchers [4, 9]; or ii) heterogeneity in the population exists and is unaccounted for, such that certain persons are at higher risk of becoming exposed or, if they are exposed, of becoming infected upon exposure for reasons other than their vaccine status, for example due to age, history of infection or vaccination, or occupation[6-8].

If either or both of these conditions hold, then over the course of the season, there will be unobserved reductions in the population at risk (or, for the second, at high risk) in each arm of the trial, and these reductions will be greater in any group that receives less vaccine protection, more moderate in a group that is more protected. In a classic comparison of vaccinated vs. unvaccinated persons, this “depletion of susceptibles” will reduce the pool of susceptible individuals (and especially of highly susceptible individuals) in the unvaccinated group more than in the vaccinated group, reducing the influenza incidence rate in the unvaccinated group relative to the vaccinated group as time progresses; equivalently, the benefit of the vaccine will appear to wane.

Recently, a novel, cohort variant of the test-negative design, was proposed and implemented that sought to circumvent these sources of bias. This design [10] considered only persons who received influenza vaccine and were subsequently tested for influenza infection. As in the classic TND the vaccine history was compared between those testing positive vs. negative for influenza infection, but unlike a classic TND, the time from vaccination to influenza test was the exposure of interest (as the study was limited to those who had received vaccine and later received a test). Relative VE for individuals vaccinated at different time points was estimated as a function of this interval, by estimating – at a specific calendar time (using conditional logistic regression) the odds ratio between influenza test-positive and test-negative participants, as predicted by time of vaccine receipt and other covariates. Crucial to this method is that individuals with different vaccination dates are compared on a fixed calendar date, rather than (as in the classic TND) comparing individuals with a different vaccination statuses on different calendar dates. The time from exposure (vaccination) to outcome (infection) is thus measured precisely and not conflated with calendar time. That study estimated approximately 16% waning in relative effectiveness of vaccination for each 28 days earlier a person had been vaccinated [10].

Peer review and a commentary published alongside the study [11] questioned whether this design had eliminated the potential bias associated with depletion of susceptibles. Subsequent discussions led to reanalysis of the data set with restriction to those who had been vaccinated before influenza season, that is, before infections with influenza could differentially deplete susceptible hosts from different time-of-vaccination groups. The result confirmed the finding of the previous analysis [12]. It was shown heuristically and with simulations that the following was true of the revised analysis: under the null hypothesis that vaccine efficacy did not wane, the study would in expectation be unbiased, estimating that indeed there was no waning, or equivalently that vaccine effectiveness was equal regardless of the time since vaccination. Under the alternative hypothesis that vaccine protection does wane, simulations showed that differential depletion of susceptibles can bias this analysis toward underestimating waning, but not toward overestimating waning and not toward an incorrect finding that VE wanes. By this logic, a study of pre-season vaccinees only that found no waning might be hard to interpret (either truly null, or waning does occur but bias in the design makes it hard to detect), but a finding that waning does occur could not be attributed to these sources of bias.

It would be ethical and informative to undertake a randomized controlled trial in which persons intending to be vaccinated are randomized to early or late vaccination, on dates anticipated to precede the start of influenza circulation (eg. September 1 vs. October 15) and incidence rates or proportions compared between these two arms, as we have proposed elsewhere [12, 13]. Knowing the expected outcomes under various scenarios would facilitate interpretation of such a trial. Meanwhile, it would be valuable to know precisely under what circumstances designs such as the test-negative case-control approach or a cohort-based modification of that approach (as performed in the example described above [10]) would perform in similar ways. For our purposes, a key difference between the classic test-negative case-control design and a prospective observational or randomized cohort design is that the latter designs attempt to track who is at risk for the outcome, for example by censoring people after they have had one influenza test [10] or after they have had one positive test (a typical randomized trial). By contrast, the test-negative case-control design relies on assumptions that the test-negative participants are representative of the population at risk. Because the biases considered in this study come from the unobserved changes in the susceptibility of the at-risk population, these may be subtly different in the different designs, and we consider several different incidence measures below that represent different approaches to tracking who is at risk.

Here, we consider a hypothetical comparison of two groups of persons, those vaccinated early (group E) and those vaccinated later (group L) with the same vaccine. These might be the two arms of a randomized trial, or might represent an idealized comparison in an observational study. When we compare two groups vaccinated at different times, with the possibility of waning, it becomes interesting to consider how either the earlier vaccinees or the later vaccinees can be subject to greater depletion of susceptibles, and thus the bias in estimating waning can go either way. Specifically, if influenza is circulating between the time when group E is vaccinated and the later time when group L is vaccinated, group L may be more depleted by incidence of infection prior to vaccination in that interval. On the other hand, if vaccine protection in fact wanes, then group E may be less protected than group L on some or all days after both groups have been vaccinated because the protection in group E will have had longer to wane. Thus, in such a scenario – where group L was vaccinated during the influenza season -- either group can be get depleted of its susceptibles faster than the other and so the bias may go in either direction. Here we show how this tradeoff occurs, and define a condition under which the bias will overstate waning (group E will look less protected than they are, because susceptibility is more depleted in group L), or will understate waning (group E will look more protected than they are, because their susceptibles are less depleted than group L), or the estimate of waning will be correct. As particular cases, we show that if vaccination of some individuals occurs after influenza season begins, and there is no waning, then the study will erroneously infer waning has occurred as a result of unobserved differential depletion of susceptibles between early- and late-vaccinated participants. If there is waning, the estimated extent of waning may be biased in either direction. On the other hand, if individuals are all vaccinated before influenza season starts (so that there is no risk of infection in any participant before they are vaccinated), and if there is no waning, the study will correctly infer that there is no waning (unbiased estimate). If individuals are all vaccinated before influenza season starts, and there is waning, then the degree of waning will be underestimated (and we cannot rule out an erroneous estimate of increased effectiveness with time since vaccination). These results are summarized in Table 1.

**Table 1:** Summary of findings

|  |
| --- |
| Truth |

Table 1: Summary of findings
|  |  | No true waning<br>(Null hypothesis) | True Waning<br>(Alternative hypothesis) |
| --- | --- | --- | --- |
|  | Include persons vaccinated after influenza season begins | Biased away from the null: Waning erroneously inferred.<br>Claim 2(i) | Biased: Waning may be over- or under-estimated depending on balance of two effects. Claim 2(ii) |
|  | Restrict analysis to persons vaccinated before start of season | Unbiased: No waning inferred (Claim 2(iii)) | Waning under-estimated. Cannot rule out erroneous rise in VE with time since vaccination (“antiwaning”).<br>Claim 2(iii)) |

## MODEL

We consider a cohort split into groups and subgroups as described below, and describe its progress through an influenza season. We define a season as a period with nonzero influenza incidence, that is the period during the year during which *λ*(*t*) > 0, where *λ*(*t*) is the force of infection with influenza, described more fully below. We denote the start of influenza season as *t*_0_. We assume that within a season it is possible to be infected with influenza at most once due to immunity. We focus on a comparison between groups with two different dates of vaccination, early (*E* vaccinated at time *t*_*E*_) and late (*L* vaccinated at time *t*_*L*_ > *t*_*E*_). We consider different scenarios where vaccination of these individuals is complete before (*t*_*L*_ < *t*_0_), or not complete before (*t*_*L*_ > *t*_0_), the start of influenza season. We envision a study in which at some time before influenza season, persons are randomized to be vaccinated early or late, or else choose their vaccination date in a way that is not confounded by predictors of the outcome (test-positive influenza). We focus here on the control of bias from differential depletion of susceptibles. In this study, all participants are vaccinated; the only difference is when. Throughout the analysis we describe expected outcomes, or equivalently outcomes in an arbitrarily large study, neglecting sampling variation; we also neglect all complexities such as loss to follow up, nonadherence, and the like, to focus on the best-case scenario for inference on the existence or nonexistence of waning. Waning is inferred to have occurred by time *t* if the incidence at time *t* is greater in the early than in the late-vaccinated group, or equivalently, the relative efficacy of the vaccine is greater in the late-vaccinated group than in the early. Note that the exact timing when the vaccine began to lose efficacy is not specified; waning may have begun before or during the season, as we infer only that it occurred between vaccination and time *t*. Note also that this definition restricts attention to *host* biological processes by which an individual’s protection from the vaccine on a given day (with the strains circulating then) is less if vaccination occurred longer ago. We define incidence in three alternative ways below, corresponding to three possible targets for estimation in different observational or randomized study designs.

Now, consider a population group *G* (this will take the value either *E* or *L* for early or late vaccinees respectively). *G* is further split into *N* subgroups of homogeneous exposure to infection and baseline “frailty” (probability of infection given exposure to infection if unvaccinated) (*i* = 1, . ., *N*) such that subgroup *G*_*i*_ is a proportion *f*_*i*_ of the population in *G*. Because we envision a large study with no confounding (by randomization or simply by assumption), the *f*_*i*_ are the same for both groups (*E* and *L*). Let *b*_*i*_*λ*(*t*) be the force of infection to unvaccinated individuals still at risk of infection subgroup *i* at time *t*. We refer to *b*_*i*_ as the frailty of group *i*, and we arrange the groups in decreasing order of frailty so that *b*_*i*_ > *b*_*i*+1_. Without loss of generality, we define *b*_*i*_ = 1. We allow for the possibility that some persons are completely immune to influenza infection throughout the season and assign them (if they exist) to the lowest-frailty group (group *G*_*N*_ with a frailty of *b*_*N*_ =0. Let *ϑ*_*G*_(*t*) be 1 minus vaccine efficacy in group *G* at time t (thus *ϑ_G_*(*t*) = 1 if *t* < *t*_*G*_, where *t*_*G*_ is the time of vaccination in group *G* and *ϑ*_*G*_ (*t*) ≤ 1after vaccination, that is when *t* > *t*_*G*_. Thus we assume the vaccine never harms an individual; it is at worst ineffective under extreme waning. For simplicity we assume that *ϑ*_*G*_(*t*_*G*_) = *ϑ*_*G*_ < 1 and *ϑ*_*G*_(*t*) is nondecreasing with *t* and is constant in the case of no waning. Thus, we assume vaccine is most protective immediately after vaccination, and may wane thereafter. Here we define waning to mean a scenario in which on a particular day, an individual vaccinated longer ago is less protected against infection with the currently circulating strains than had they been vaccinated more recently. We assume that vaccine efficacy, and equivalently *ϑ*_*G*_(*t*), is the same for all subgroups *G*_*i*_ within *G*; this assumption may be loosened but is kept for the sake of clearer exposition in the proofs.

Let 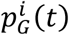 be the proportion of persons in subgroup *G*_*i*_ still at risk of influenza infection at time *t*. Because we have placed all persons totally immune to infection in group *N* with frailty *b*_*N*_ = 0, we can assume that everyone in groups with nonzero frailty is susceptible at the start of flu season, that is, 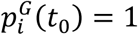 if *b*_*i*_ > 0.

The proportion at risk in group *G* as a whole is

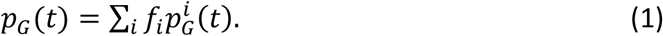

For each subgroup *i*, rate of change with time is

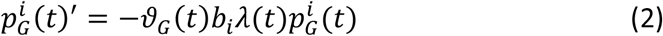

We define the mean frailty among those still at risk in group *G* as

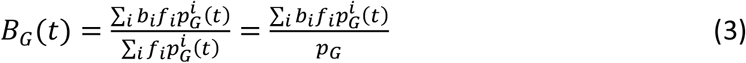

If a proportion *a* of all cases is ascertained (ie is symptomatic and comes for testing and tests positive for influenza), then the rate at which influenza cases in group *G* present for care and test positive for influenza, following the notation in ref. [4], but dropping the subscript for influenza, is

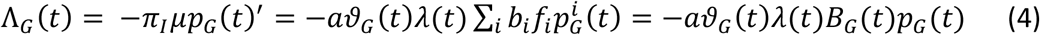

We note that the proportion of the population at risk in each group at time *t*, which we call *p*_*G*_(*t*), will in general differ from the proportion the investigators believe to be at risk in that group, as long as not all cases are ascertained [9]. The proportion thought to be no longer at risk will be the cumulative number infected, times the probability of ascertainment given infection. Denoting this probability of ascertainment as *a* = *π*_*I*_*μ*, the proportion thought to be at risk in group *G* at time *t* is

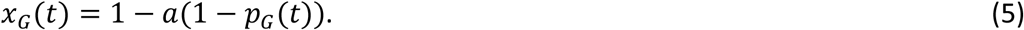

## RESULTS

The following claims state formally the conclusions summarized in Table 1. We describe each claim, and give the proofs in the appendix.

### Claim 1

Suppose that the influenza season begins at time *t*_0_ after which influenza hazard of infection *λ*(*s*) ≥ 0 for *s* > *t*_0_, and let the early and late groups be vaccinated at times *t*_*E*_ and *t*_*L*_ respectively. These may be before or after *t*_0_. We consider various incidence measures at time *t*_1_ > max(*t*_*L*_, *t*_0_). Whenever the cumulative hazard for the highest-frailty subgroup, modified by vaccination, by time *t*_1_ in group E is less than in group L, which is equivalent to

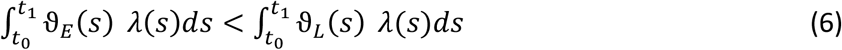

we will have

a. 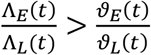
b. 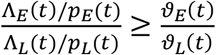
c. 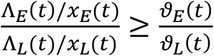 and
d. if inequality 6 is reversed, then inequalities a, b, and c are reversed. Inequality b will be strict if there is heterogeneous frailty (*N* > 1). Inequality c will be strict if there is heterogeneous frailty (N>1) and/or imperfect ascertainment of cases (*a* < 1), and equal otherwise (*a* = *N* = 1). All inequalities will become equalities if the two sides of eq. 6 are equal.

Remark: this claim concerns three different incidence measures that may be of interest.

a. 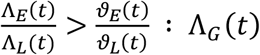 is the incidence measure treating the original population at risk as the denominator.
b. 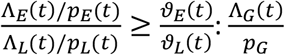 is an incidence measure including in the denominator only those participants in the denominator who have not yet been infected at *t*. Here the inequality is strict if there is heterogeneous frailty, but if frailty is homogeneous (only *N* = 1 subgroup in each group) then equality holds and no waning would be inferred.
c. 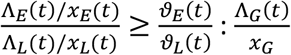 is incidence among those who were at risk at the start of the season and are not known to have been infected before *t* (allowing for imperfect ascertainment of each case with probability *a* as defined in equation (5).

### Claim 2: The particular cases considered in Table 1 are true, following from Claim 1

i) top row of Table 1: If there is no waning (so that 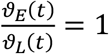 for *t* > *t*_*L*_) and vaccination is not completed before the start of influenza season (*t*_*L*_ > *t*_0_), then for all for *t* > *t*_*L*_, the following inequalities will hold, potentially producing erroneous inferences of waning:
  a) Λ_*E*_ (*t*) > Λ_*L*_(*t*)
  b) 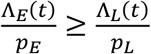: Here the inequality is strict if there is heterogeneous frailty, but if frailty is homogeneous (only *N* = 1 subgroup in each group) then equality holds and no waning would be inferred.
  c) 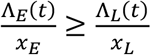. Here, the inequality is strict if there is either heterogeneous frailty (*N* > 1) or imperfect ascertainment (*a* < 1), but equality holds if neither of these applies (*a* = *N* = 1).
ii) If there is waning and vaccination is not completed before the influenza season, the net bias may go either way. If equation 6 holds and 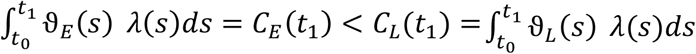, then waning will be overestimated, but if the inequality is switched, it will be underestimated.
iii) (bottom row of Table 1): If vaccination is completed before influenza season begins (*t*_*E*_ < *t*_*L*_ < *t*_0_), then the following inequalities will hold, with waning underestimated when it exists and correctly estimated as null when it does not.
  a) 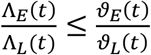with equality under the null of no waning 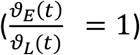
  b) 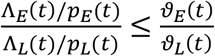 with equality under the null of no waning or when frailty is homogeneous (*N* = 1)
  c) 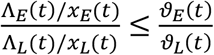 with equality under the null of no waning or when *a* = *N* = 1, ie both (i) frailty is homogeneous and (ii) case ascertainment is perfect.

## DISCUSSION

We have formalized and proved in the appendix the claims summarized in Table 1 about the direction of bias when various study designs are employed to assess whether vaccine protection against influenza infection wanes within a season with increasing time since vaccination. If a study compares the incidence of influenza among persons with early vs. late vaccination, and if all vaccinations are completed before the start of influenza season, the design will be unbiased under the null: no waning will be inferred. Under the alternative hypothesis that waning does occur, its extent will be underestimated. Therefore, if waning is inferred, the inference that it is occurring is robust, and the true magnitude may be larger than what is inferred. It is theoretically possible that early vaccination could look more protective than late under such a scenario when comparing instantaneous incidence because of the phenomenon of crossing hazards [4, 14], though the practical likelihood of such intense bias may be small. On the other hand, in a design where some vaccinations occur after the start of influenza season, the estimate is biased under the null: if there is no waning of vaccine-induced protection, waning will be inferred spuriously. If there is waning, the direction of bias is not determined.

The demonstrations of each of our findings for the vaccinee-only design rely on the same principle, applied differently when the timing of vaccination relative to the season is different. The common principle is that a group that has more vaccine-induced protection will retain a higher proportion of susceptible or highly susceptible individuals, while these will be depleted faster in the group with less vaccine-induced protection. The investigators will be unable to track this differential depletion if (1) susceptibility (frailty as we called it in line with other literature) is variable but unmeasured and/or (2) infections are not all ascertained (eg due to some being mild), so the population at risk is less than that thought to be at risk, especially in the less-protected group.

This common principle is applied in opposite ways in different scenarios, because the late-vaccinated group is more depleted when some influenza incidence occurs before they are vaccinated, and the early-vaccinated group is more depleted when protection wanes, making them less protected. In claim 1, we show how these alternative directions of bias balance when both are present, with bias toward less waning if the effect of waning dominates, and bias toward more waning if the depletion of susceptibles from the late-vaccinated group before they received vaccine dominates. In claim 2, we apply this to particular cases and mathematically confirm previous heuristic results – that waning estimates would be null when there is really no waning if vaccination is complete before influenza circulation, that waning would be underestimated if it truly exists and vaccination is complete before influenza circulation, and that waning will be erroneously inferred if it does not exist if vaccination is incomplete at the start of the influenza season. These lead to the recommendation to restrict waning studies to persons vaccinated before influenza season begins.

Estimation biases occurring due to cohort-selection, differential depletion of susceptibles, or unaccounted-for frailty heterogeneity (three terms for the same phenomenon [4, 6, 8, 9, 14, 15]) have been recognized in the literature for some decades[4, 6, 8, 9, 14, 15] but are often not accounted for in study design and analysis. The analysis here contributes two aspects to the discussion. First, it mathematically separates out the effect of heterogeneous frailty (variation in *b*_*i*_ in our notation, emphasized for example in (in review) and [7, 16]), which leads to the less-protected group being more rapidly depleted of its most frail members and thus looking less at-risk in the aggregate, from the effect of having unobserved infections (more of these in the less protected groups) that deplete the number of persons at any risk differentially from different groups, emphasized for example in [4, 9]. These biases work in the same direction, so that the biases discussed here arise when either or both are present. The second contribution is to show a general condition under which biases in one direction or the other are dominant in a comparison of persons vaccinated on two dates, depending on which group has been more depleted of susceptibles. The third is to show in general that, as proposed in (manuscript in review), designs that restrict comparison to times of vaccination before the onset of disease exposure are not susceptible to spurious inference of waning. While not applicable in all cases [7], this may be achieved conveniently in highly seasonal diseases where a vaccine can be delivered before transmission begins – such as influenza in temperate climates.

We note that this analysis considers only the biases that result from susceptible depletion (which can be seen as a form of selection bias [17]). It does not consider other issues of confounding and selection bias that can plague observational studies in this area [18, 19]. Therefore, it is notable that these concerns apply even in randomized trials; the reason can be clearly seen, in that the biases occur due to post-randomization differences that arise between the two arms and influence the outcome (incidence). The exact degree of the bias depends on details of the study design, however. We showed that a bias in the same direction occurs for each of three incidence measures. The first (daily rate of reported cases, without reference to a population at risk) would be most relevant to the classic test-negative case-control design, where no explicit cohort is followed (so depletion of susceptibles is entirely unobserved) but rather, incidence of “test-negative” infections is used to assess the population at risk indirectly. The last (rate of reported cases, relative to a population at risk that has been reduced when cases are observed (since by assumption no one can get influenza twice in a season) is most relevant to a randomized controlled trial or a study similar to that of [10], where a cohort is followed, and persons receiving an influenza diagnosis are removed fro the at-risk group (this particular study also removed those who received an influenza test and were negative, but this does not change the general finding). The middle incidence measure would be a target for estimation in a study where every influenza case would be diagnosed and removed from the at-risk group [9]. We considered this to make explicit that, even if this is accomplished (eg by virologic or serologic testing [9]) the existence of variable frailty will still lead to the bias. Only if frailty is homogeneous and all infections are perfectly ascertained (or if the vaccine is entirely ineffective, perhaps due to a mismatch) does it completely disappear in general [9]. In the special case where there is no waning, however, the design with preseason vaccination only will be unbiased, and if there is waning, the preseason vaccination design will not overestimate its extent. Therefore a finding of waning under that design (as in (under review)), is compelling (unless other important biases are posited), while a failure to detect waning with that design is harder to interpret.

In summary, we have provided evidence that a small modification to some existing studies of vaccine waning – specifically, restricting consideration to those vaccinated before influenza season -- may be sufficient to make findings of measurable waning very convincing and worthy of consideration in recommendations for the timing of vaccination. We recommend such an approach in future studies, whether experimental or observational.

## Data Availability

not applicable

## ACKNOWLEDGMENTS, FUNDING AND COMPETING INTERESTS

Support for this work came from: the National Institute of Allergy and Infectious Diseases, National Institute of Health, USA grant #1R01AI107721-01; cooperative agreement U54GM088558 from the National Institute Of General Medical Sciences, National Institutes of Health USA; UK National Institute for Health Research (NIHR)grant PR-OD-1017-20006 (Epidemiology for Vaccinology stream) using UK aid from the UK government; and The Permanente Medical Group. The content is solely the responsibility of the authors and does not necessarily represent the official views of the National Institute of General Medical Sciences, the National Institutes of Health, the NIHR or the UK Department of Health and Social Care. G. Thomas Ray reports research support from Pfizer. Marc Lipsitch reports support from Merck, Pfizer, Antigen Discovery, and Affinivax. The remaining authors have no potential conflicts of interest to disclose.

## Appendix

### A. A useful result applied in the main proof

#### Generalized Grönwall inequality

Suppose we have two functions *f*(*t*), *g*(*t*) that are solutions to the following ODEs

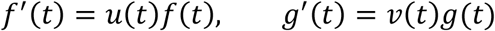

If *f*(0) ≥ *g*(0) > 0, one also has that *f*(*t*) ≥ *g*(*t*) for *t* > *t*_0_ as long as

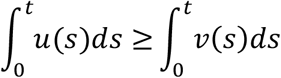

In particular, that holds if *u*(*s*) ≥ *ν*(*s*).

Moreover, the inequality *f*(*t*) ≥ *g*(*t*) is strict if either *f*(0) > *g*(0) or 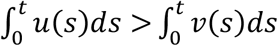

**Proof:** The ODE for *f*(*t*) can be re-written as 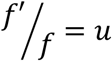, which mean 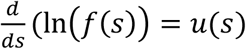.

Integrating this from *t*_0_ to *t* we get that

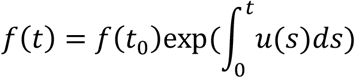

similarly,

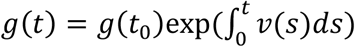

From this the Generalized Gronwall Equality follows.

### B. Proofs of the main claims

#### Proof of claim 1

By eq. 4,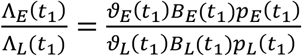. We prove here that when eq. 6 is true,

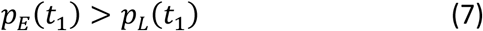

and

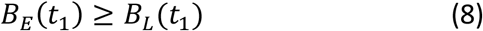

Together these demonstrate claim 1a, and eq. 8 alone demonstrates Claim 1b.

Proof that eq. 6 implies eq. 7:

For each *i*, eq. 6 implies

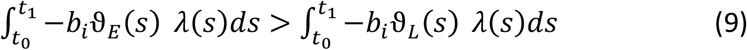

Using eq. 3, let 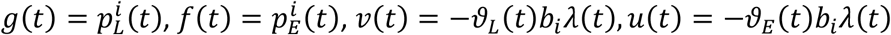 in the notation of the Generalized Gronwall Inequality. Eq. 9 then satisfies the condition of the Generalized Gronwall Inequality. This implies that 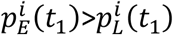, for all *i* and thus by eq. 1 that *p*_*E*_(*t*_1_) > *p*_*L*_(*t*_1_). This is eq. 7, QED

Proof of eq. 8 when eq. 6 holds:

Assume there are at least two subgroups with different frailties: for subgroups *i* and *j*, with *i* < *j*, we will have *b*_*i*_ > *b*_*j*_. Then eq. 6 implies eq. 9, which implies for these two subgroups:

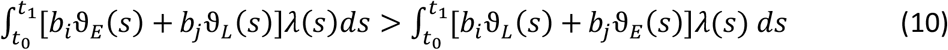

Now, to show that *B*_*E*_(*t*_1_) < *B*_*L*_(*t*_1_), we need to prove that at time *t*_1_

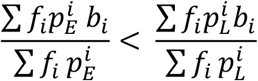

Subtracting the l.h.s. from the r.h.s. we get

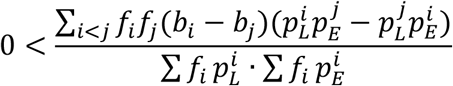

Recall that we ordered *b*_*k*_ in the descending order so that *b*_*i*_ > *b*_*j*_ when *i* < *j*. We need, for *i* < *j*, and *t* > *t*_0_ to show that

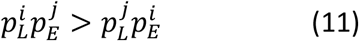

We note that the two sides of eq. 11 are equal at *t* = *t*_0_

Differentiating the function 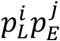 and using the Generalized Gronwall inequality, we note that this function is a solution to the ODE

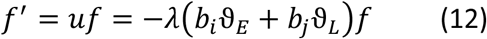

Similarly, the function 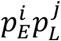 is a solution to the ODE

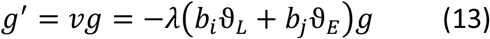

Thus eq. 8 will hold when

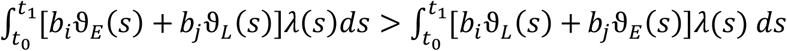, but this is eq 10, so we have proven that eq. 6 implies eq. 8.

Note that the foregoing relied on heterogeneous frailty ((more than one group with different values of *b*_*i*_). When there is one level of frailty (*N* = 1, *b*_1_ = 1), *B*_*G*_(*t*) = 1 for all *G, t*.

Having proven eq (7) and eq (8) we have claim 1(a), with always a strict inequality. Having proven eq (8) alone we have claim 1(b). The inequality is strict when there is more than one subgroup with different frailties; otherwise, we have equality.

#### Proof of claim 1(c)

To show that 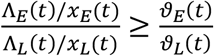, when eq. 6 holds, we note that we have proven 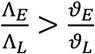 and 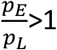 when eq. 6 holds. But 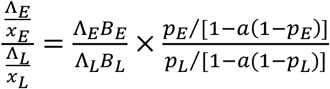. Given that *a* ≤ 1 and *p*_*E*_ < *p*_*L*_ ∈ (0,1], a little algebra shows that that 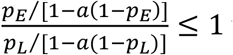 with equality when *a* = 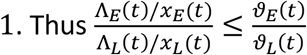 with strict inequality either *a* < 1 (imperfect ascertainment, making the *x*_*G*_ inequality strict) or *N* > 1 (heterogeneous frailty, making the *B*_*G*_ inequality strict. We have equality for claim 1(c) when *a* = *N* = 1.

#### Proof of claim 1(d)

All of the foregoing proofs are symmetric in groups E and L. If 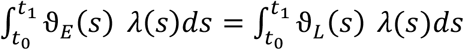 then results 1(a)-(c) hold with the inequalities reversed, proven by identical arguments. Likewise, if the two sides are equal, then all quantities in the proofs will be equal between groups and claims 1a-c will be show equality.

#### Proof of claim 2(i)

If there is no waning (so that 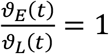 for *t* > *t*) and vaccination is not completed before the start of influenza season (*t*_*L*_ > *t*_0_), then 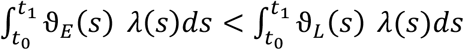 because group E will experience protection (ϑ_*E*_ (*s*) < 1 = ϑ_*L*_(*s*)) for the time between the start of the season or vaccination in group E (whichever is latest), and vaccination of group L (max(*t*_0_, *t*_*E*_) < *s* < *t*_*L*_), and thereafter ϑ_*E*_(*s*) = ϑ_*L*_(*s*) = *ϑ*. Therefore the condition of Claim 1 is fulfilled, so

By Claim 1,

a) 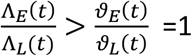
b) 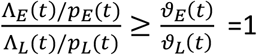
c) 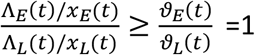

In the case of (b), there is equality when frailty is homogeneous, and the inequality is strict when there is heterogeneous frailty (more than one group with different values of *b*_*i*_), as noted in Claim 1(b). In the case of (c), there is equality when ascertainment is perfect and frailty is homogeneous (*a* = *N* = 1), and strict inequality otherwise.

#### Proof of claim 2(ii) (Top right of Table 1)

If vaccination is incomplete at the start of the influenza season and waning occurs, then there will be conflicting biases due to depletion of susceptibles. Rearranging eq. 6 we have:

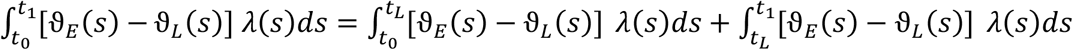

where the first integral on the right is negative due to earlier vaccination of group E, and the second integral is positive due to waning. The balance determines whether the extent of waning will be overestimated or underestimated.

#### Proof of claim 2 (iii) (bottom row of Table 1)

If vaccination is completed before influenza season begins (*t*_*E*_ < *t*_*L*_ < *t*_0_), then the following inequalities will hold, with waning underestimated when it exists and correctly estimated as null when it does not. This comes from an application of Claim 1, with the sign reversed (if there is waning) or equality (if there is no waning). If vaccination is complete before influenza season, then the only source of differences in 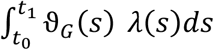 is waning; otherwise the cumulative vaccine-adjusted incidence will be equal between groups throughout the study, which will give 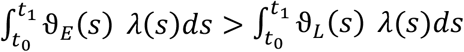. Therefore the condition of Claim 1 is satisfied (with the inequality reversed) if there is waning, and equality holds in the condition of claim 1 under the null of no waning. From this it immediately follows that:

a. 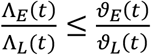 with equality under the null of no waning 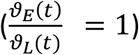
b. 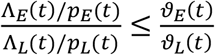 with equality under the null of no waning or when frailty is homogeneous (*N* = 1)
c. 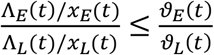 with equality under the null of no waning or when *a* = *N* = 1, ie both (i) frailty is homogeneous and (ii) case ascertainment is perfect.

